# SEDRA: Selective Entry Dynamic Risk Assessment: A mathematical model to safely keep the borders open during Covid-19

**DOI:** 10.1101/2021.12.05.21267311

**Authors:** Silvano de Gennaro, Håkan Lane

## Abstract

The Covid-19 pandemic has brought the World to a near standstill for most of 2020 and 2021, causing chaos in international travel, driving many economies into the ground, particularly those largely based on tourism. The lack of standard tools to assist decision makers in structuring a coherent policy to allow foreign passengers into their county and the resulting panic-mode opening/closing the borders on every “new case” outburst or new variant “of concern”, have led several countries to costly and often meaningless decisions based on fear rather than science or logic. This study aims at providing a universal method to safely keep the borders open and allow conditional immigration to foreign passengers according to a “Risk Group” table that includes all the countries reporting data on their Covid-19 situation to the WHO and other organisms. The RG table is recalculated on a weekly basis according to a mathematical model described in this paper, dynamically assessing the status of the pandemic worldwide through the calculation of a “Safety Index” for each country. A prototype algorithm has been implemented in VBA/EXCEL and its results are published bi-weekly on a Github repository.

## 2. Introduction

The Covid-19 pandemic has paralyzed the World’s economy over the last 2 years, by leading many countries to a series of national and regional lockdowns as well as major restrictions on flights and human mobility, penalizing many sectors and mostly those related to tourism and air travel. This study intends to provide a mathematical model that that can help countries with the regulatory conditions for reopening their borders to tourism and immigration in general. The method consists in calculating a “*Safety Index*”, namely a number ranging from 0 to 100, raking the safety level of each country as a source of Covid-19 contamination. The Safety Index is calculated as an inverse function of the risk associated to a given country for each arriving passenger and it is based on the following parameters:

a. The probability that a passenger arriving from a given country is infected despite testing negative (undetected infective)
b. The mortality registered in the last week in that country
c. The confidence level of the data submitted by that country

The first two parameters can be considered as indicators of the status of the virus transmission and severity, while the third one tells us how reliable the first two are. The formula relating these three parameters will be described in the next chapter.

This study takes into account the following considerations

1. The situation of the pandemic may change rapidly from a week to the next, therefore the risks must be reassessed, i.e. recalculated on a weekly basis.
2. All calculations must be performed using *relative* numbers rather than *absolute*. A relative number is the number of cases, tests and deaths per million inhabitants.
3. The maximum incubation period of the SARS-CoV-2 virus was initially of 14 days but has lowered considerably with the arrival of the current dominant variant, omicron. During this period the great majority of infected persons could produce a negative PCR^1^ test.

Ultimately, the Safety Index will allow us to assign each country to a “Risk Group” (RG) ranging from A (Very Low) to E (Very High). It will ultimately pertain to governments to decide upon the restrictive measures to apply for each RG, however. For instance, free entry for groups A and B, PCR test for group C, 7 days self isolation for group D and quarantine for group E. The attribution of countries to RGs, based on the calculated Safety Index is done through the setting of thresholds as in the following example:

The choice of expressing the Safety Index as an integer number in the interval [0, 100] rather than a probability is justified by making it easily understandable to the large public and decision makers, often unfamiliar with mathematical concepts and infinitesimal fractional numbers requiring an exponential notation.

## 3. Safety Index calculation

This chapter covers the mathematical model used for the calculation of the Safety Index. Readers unfamiliar with algebra or not interested in the details may skip to chapter 4, where we will resume the main concept of the model in a non-mathematical way.

We use the data series published by *OurWorldinData* (4). We refer to the countries in that table as a series *C*_*i i*=1..*n*_. For each country we define the following corresponding parameters:

- ***Immunity Level*** (*Y*_*i*_): an indicator in the interval [1,10] of the probability that a passenger coming from country *C*_*i*_ is **not** infected
- ***Confidence Level*** (*K*_*i*_): an indicator in the interval [1,10] of the reliability of the data reported by country *C*_*i*_.
- ***Severity Level*** (*S*_*i*_): an indicator in the interval [1,10] of the relative death toll in country *C*_*i*_.

We will then define the ***Safety Index*** (*Z*_*i*_) as a function of the above parameters:

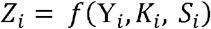

### Probability of undetected infection

We start by calculating the probability of arrival of an undetected infective (UI) from country *C*_*i*_. In order to be infected and still test negative at arrival, a passenger must be still in the incubation phase. This is a variable number, depending on the currently dominant variant. It has been showed that this figure was 14 days for the previous variants (alpha to delta) but it has greatly reduced with the omicron BA.4/BA.5 (max: 5 days, µ: 3.5) [6]. We shall use the symbol ℑ to express the incubation period (days).

A passenger in his/her incubation phase must have been in contact with an individual who has been infected in turn within the past ℑ days. Although it is impossible to know the exact number of new infections in a country, we can assume that it is proportional to the new confirmed positive cases in the last ℑ days.

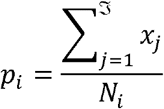

Where:

*p*_*i*_ = Probability of a positive case arriving from country *C*_*i*_

*x*_*j*_ = New daily positive cases in the last ℑ days (*j* = 1.. ℑ) in country *C*_*i*_

*N*_*i*_= Population size of country *C*_*i*_

ℑ = Max incubation time for current dominant variant

This probability is in most countries a very small number in the order 10^−3^ – 10^−7^ and its smallest realistic nonzero value corresponds to the probability that one single person is currently an undetected infective in the largest country in the World, China. That is :

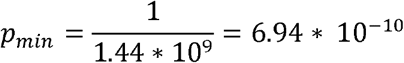

For the purpose of this study we will focus on the order of magnitude of these probability values, which is represented to a large extent on the exponent of the above result.

### Immunity Level

The Immunity Level quantifies the opposite of infection and can be expressed as the absolute value of the logarithm of the probability of infection.

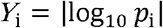

Where:

*Y*_*i*_ = Immunity level for country *i*

Immunity Levels are therefore positive real numbers in the range 0… 10. According to this formula, the Immunity Level of China as described above would be 9.15816, which corresponds to the maximum possible value on Earth for a non covid-free country. As for covid-free countries, defined as those countries that registered zero new positive cases in the last ℑ days, the value of 10 will be forced by the Risk Assessment algorithm.

### Confidence Level

The above calculation relies upon the quality of the official data provided by the WHO, the John Hopkins and similar institutions, and collected by Our World in Data on the platform: www.github.com (4). This quality depends upon the national reporting schemes. Unfortunately the accuracy of the latter differs greatly across the World and the data is often incomplete, unprecise or totally missing. Therefore in order to trust the values of *p*_*i*_ in the formula above, we need to assess its reliability for each country, and the best way to do that is to calculate a confidence level *K*_*i*_ as a function of the average daily testing per million people performed. The larger the testing sample, the more accurate the value of *p*_*i*_ will be. Once more we will evaluate this as an average over the past ℑ days, and we will normalize it to match the range of Immunity Level [1,10], by using a linear function:

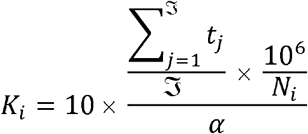

Where:

*K*_*i*_ = Confidence Level

*t*_*j*_ = Average new daily tests per million in country *C*_*i*_, over the past ℑ days

*N*_*i*_ = Population size of country *C*_*i*_

*α* = Boundary (maximum expected) value of daily tests per million worldwide

ℑ = Max incubation time for current dominant variant

Substantially we calculate the average value over the incubation period, we make it relative by dividing by the population and multiplying per one million, divide by *α*, the maximum relative testing ever done to bring it in the [0,1] range, then finally multiply by 10 to bring it in the [0,10] range, in order to match the values of the Immunity Level. At the time of writing of this paper, the value of *α* is 147, recorded by Cyprus.

This formula can be written in a more compact form as:

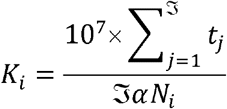

### Severity Level

Another parameter that we must take into account when deciding about opening the borders to a certain country is the severity of the disease. This can be quantified by looking at the relative average number of *daily deaths per million*. This amount can of course be modulated by new variants, by the efficiency of the heath system, by climate, viral load, stringency index, country policies and political agenda, but it is the only numerical reference we have to determine how dangerous the virus in a given country is.

We will use the same method we used to calculate the Confidence Level:

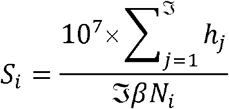

Where:

*S*_*i*_ = Severity Level:

*h*_*j*_ = new daily deaths in country *i*, with *j* =1.. ℑ days

*N*_*i*_ = Population size of country *i*

*β* = Boundary (maximum expected) value of daily deaths per million worldwide

ℑ = Max incubation time for current dominant variant

Caution: The value of *β* must be assessed realistically. In small countries, with population below 1 million, the values of “deaths per million” can be misleading and bias the scale for all other countries. For instance, the top ranking in deaths per million is the Rep. of San Marino, with 63.14 deaths per million. However the population of that country is only 33,938 and the total number of deaths is 20. On the other hand, a more realistic maximum is recorder by Ecuador, with a population of 17.6 million and a total number of deaths of 10,900, where we find a more realistic value of 33.17 smoothed new deaths per million. Therefore in order to avoid extrapolation errors, we will ignore countries with a population of < 1 million in setting boundary values. A value of 0 will be forced for *S*_*i*_ by the algorithm for these countries.

### Safety Index

We will finally relate the parameters we defined in a single formula to calculate a *Safety Index* that can assess in a single figure the risk presented by each country analyzed:

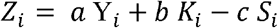

Where:

*Z*_*i*_ = Safety Index for country

*a,b,c* = weights

This formula positively binds the *Immunity Level* with Confidence and negatively with *Severity*, forcing higher values for more reliable data and lower ones for countries experiencing high death rates in the last ℑ days. The fine tuning of this formula is done through the constants *a,b,c*, (weights) which are set by the user depending on how much importance they wish to give to *Confidence* and *Severity*.

By substituting the three parameters, we obtain a compact version of the *Safety Index* formula:

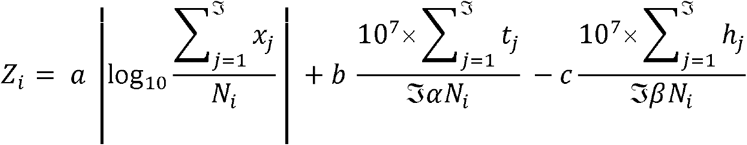

### Risk Group

We will now relate the *Safety Index* to *Risk Groups* through a threshold table that simply defines intervals for the wanted RGs. As an example, let’s suppose we want to define 5 RGs, ranging from lower to higher risk, as follows: A, B, C, D, E.

We will then create an association table using threshold values that we set manually and we will name *t*_l_, *t*_2_, *t*_3_, *t*_4_:

A (Very Low): *t*_4_ ≤ *Z*_*i*_ < 100

B (Low): *t*_3_ ≤ *Z*_*i*_ < *t*_4_

C (Medium): *t*_2_ ≤ *Z*_*i*_ < *t*_3_

D (High): *t*_1_ ≤ *Z*_*i*_ < *t*_2_

E (Very High): 0 ≤ *Z*_*i*_ < *t*_1_

As an example, with following settings:

- Weights: a = 10; b = 1; c = 3
- Thresholds: *t*_1_= 25; *t*_2_= 35; *t*_3_= 45; *t*_4_= 60

We obtain the resulting table below (Table 2) for the first 38 countries in alphabetical order, based on the OurWorldinData dataset (4) on 16/6/2022.

**Table 1:** Example of Risk Group ranking according to Safety Index thresholds

|  |  |  |
| --- | --- | --- |
| Risk Group | Risk Level | Safety Index thresholds |
| A | Very Low | $50 < SI \leq 100$ |
| B | Low | $40 < SI \leq 50$ |
| C | Medium | $30 < SI \leq 40$ |
| D | High | $20 < SI \leq 30$ |
| E | Very High | $0 \leq SI \leq 20$ |

**Table 2:**
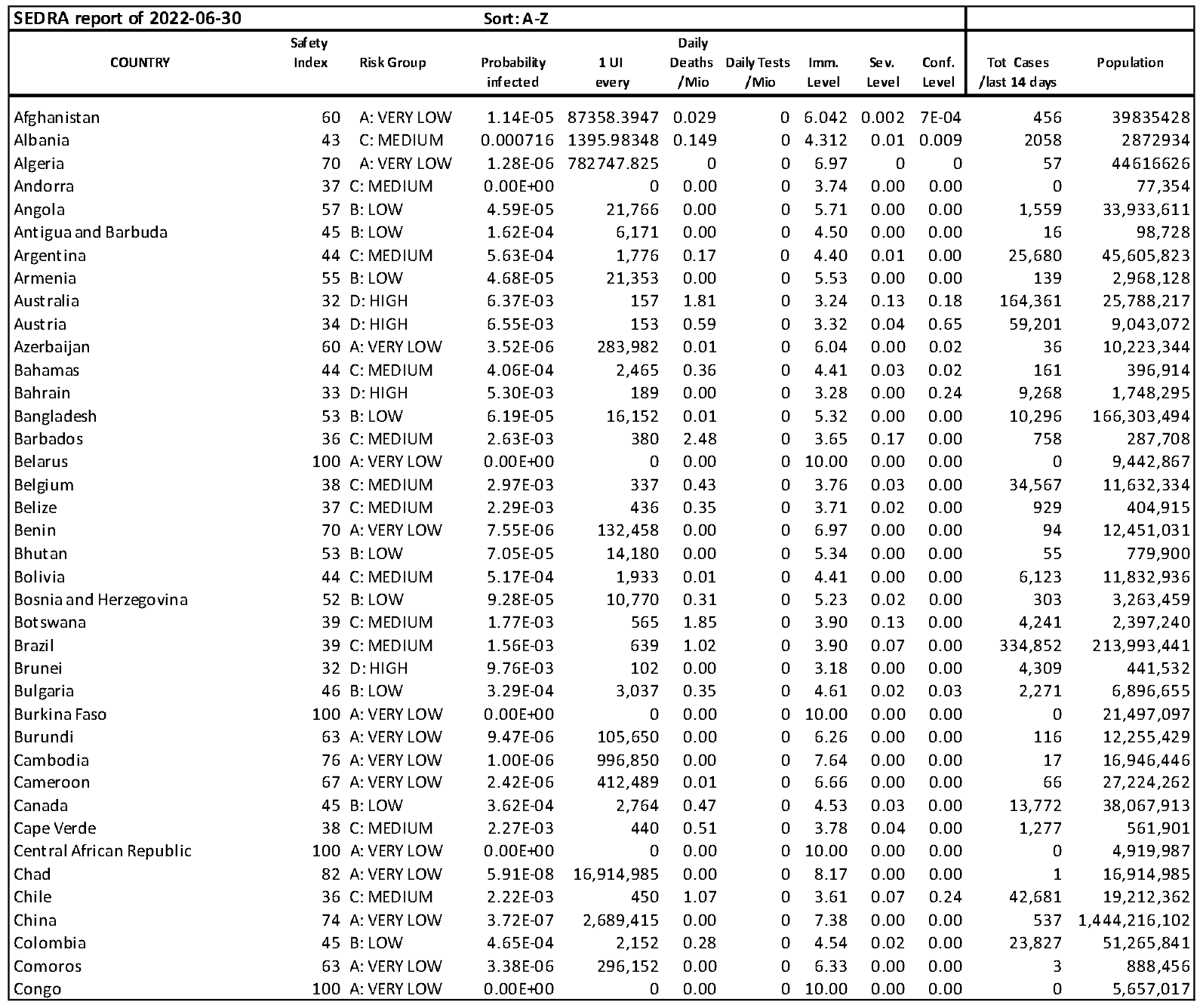
Example of risk assessment calculation. OWID data of 30/6/2022.

The parametrization of this algorithm through weights and thresholds allows us to follow a stricter or loser risk strategy:

Example of stricter risk strategy:

- Weights: a = 10; b = 3; c = 3
- Thresholds: *t*_1_ = 35; *t*_2_ = 45; *t*_3_ = 60; *t*_4_= 75

Example of loser risk strategy:

- Weights: a = 10; b = 0; c = 1
- Thresholds: *t*_1_ = 20; *t*_2_= 35; *t*_3_= 45; *t*_4_= 60

An example of SEDRA table sorted by Safety Index can be found in Appendix A.

Appendix B shows two charts for the Safety Index. Although it is difficult to follow each single line, due to the tightness of the range, these charts can help visualize at once the progression of the pandemic in a certain region. We can clearly see that the SI was generally increasing in Southern Europe with the advent of Summer (July), but it deteriorated prematurely due to the arrival of the Delta variant in August. It then grew again with the arrival of the new dominant variant omicron.

The complete repository is archived bi-weekly on: https://sedraproject.github.io/index.html

This repository contains the SEDRA Charts for all regions and continents in a interactive format, allowing to single out each country by hovering on the line graph.

## 4. Case studies

### One day snapshot

September 2, 2021, was chosen as an example of daily analysis generated by the SEDRA algorithm. According to the Worldometers data gathering site, Worldwide, there were 677804 new cases and 11265 deaths related to the SARS-Cov2 virus. Figure 1 shows the distribution of risk classes by continent.

**Figure 1.**
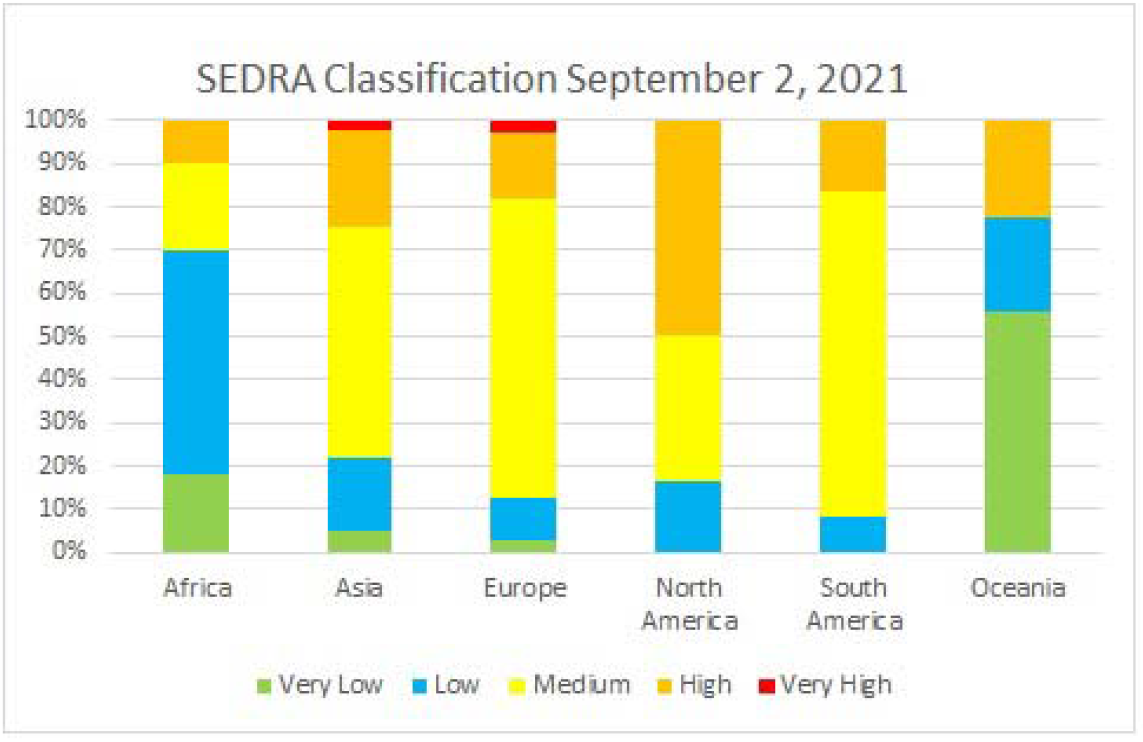
Risk classification by continent, September 2, 2021.

It is worth noting that most countries especially in Europa, Asia and the Americas were still at high or very high risks, while Africa and Oceania were relatively safe.

### Timelines

A country needs to keep borders to another nation closed really only if the risk profile is significantly higher abroad. By looking at the timeline of risks for two countries, it would be possible to estimate timeframes when international travel between those locations should be deemed safe. The risk scores for Germany and Mauritius, respectively, are indicated in Figure 2

**Figure 2.**
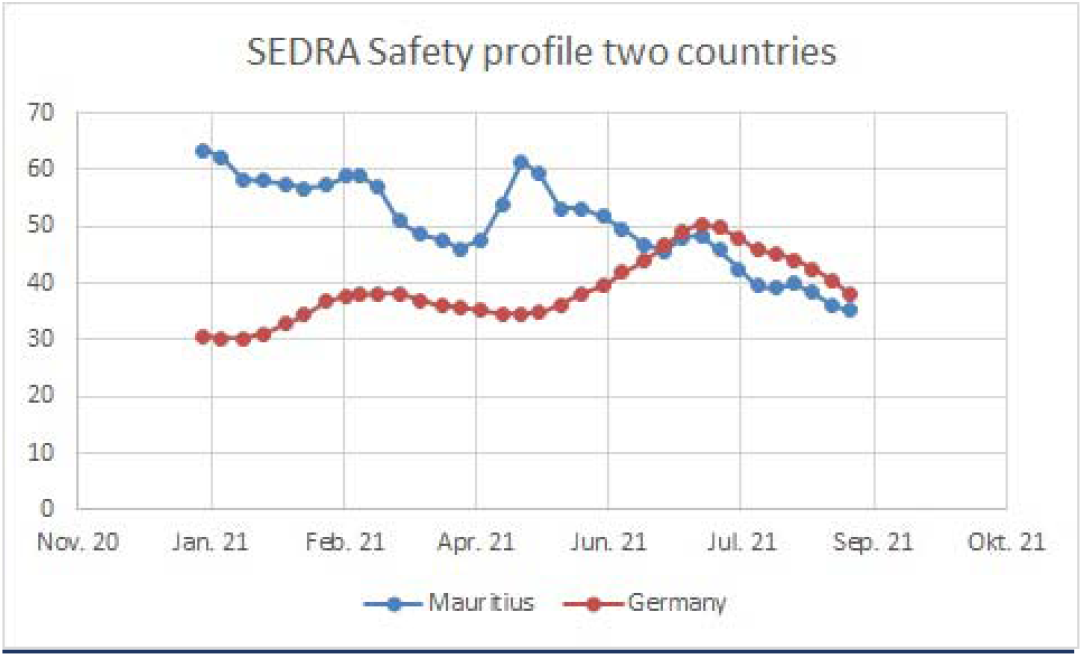
Risk values September 2, 2021.

As higher values indicate less risk, the countries were at par with eachother from mid-June and onwards, which should have made international travel possible.

### Comparison major tourism countries

Figure 3 further underlines the case for a possible sooner reopening of international travel to the example of Mauritius. The image shows the distribution of safety class on August 19, 2021, among the 16 biggest sources of tourism to the island.

**Figure 3.**
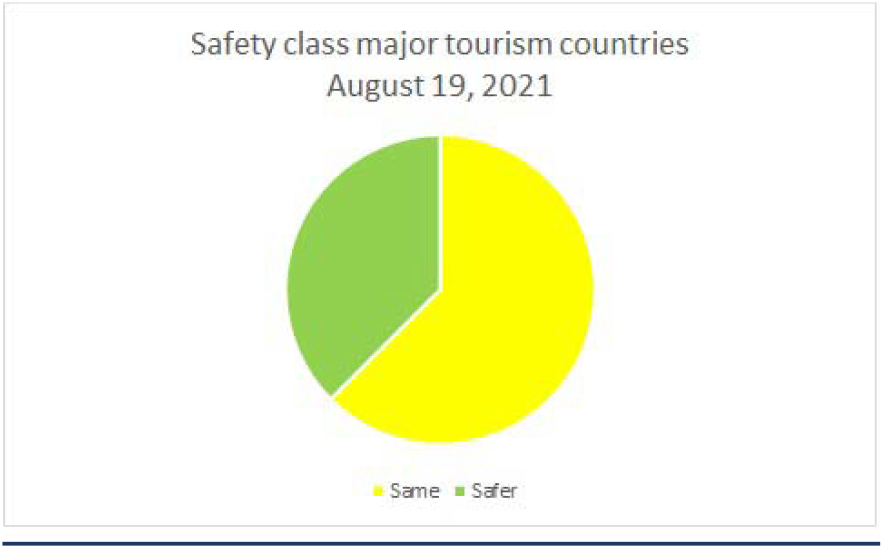
Comparison Mauritius major tourism sources August 19, 2021.

Out of the 16 most frequent sources of tourism to Mauritius, all were either in the same class or safer on August 19, 2021.

## 5. Discussion

In the previous chapter, we have developed a mathematical model that allows to bring the probability of arrivals of undetected infectives from a given country, the severity level (death rate) and confidence level within the same scale (1..10). We have then presented a simple formula to use these indicators in order to assess the risks of importing new positive Covid-19 cases from that country using a single parameter, the Safety Index.

If adopted globally, this method can allow all countries to follow a similar, consistent defense line against the propagation of the virus across the globe.

This formula does not explicitly take into account vaccinations, variants, climatic conditions, viral load in the country of origin, however those parameters are implicitly correlated to the indicators we have chosen.

### Vaccination

During 2021, many countries have lifted all restrictions for passengers carrying a WHO recognized vaccination passport. A vaccinated person is in fact believed to be safe to a certain extent from a serious evolution of the disease however he/she can still be a healthy infective of the virus in its incubation phase. In other words, the “vaccination passport” is not a guarantee that the visitor is not infected. In particular vaccinated subjects infected with the Delta variant could carry the same viral load as the unvaccinated (2).

### Variants

Some of the new variants of the virus such as omicron (BA.4/BA.5) have shown to be more contagious, more aggressive, less detectable by a PCR test and immunoresistant to both vaccines and previous infection [7]. On the other hand it shows a shorter incubation time and a lower severity, reducing mortality to a level just higher than a strong flu and calling for many countries to lift all restrictions. Virologists are however cautious and warn that it is too soon to call this the end of the pandemic [8], since new mutations with unpredictable pathogenicity are still highly likely, leading to more disruptions of World travel, economy and lifestyle.

## 6. Immigration measures

This study gives regulators and decision makers a mathematical framework to help them set immigration rules from any foreign country. It guides them towards decisions based on statistical risk assessment rather than media sensationalism. For example, until April 2022, passengers coming from many Covid-free and Risk Group A (very low risk) countries had to do quarantine when arriving in Italy. This unreasonably penalizes tourism and business since these passengers have very little chance of being infectives. But they are being quarantined simply because the country of arrival does not know how to process the data about the country of origin. This study provides that data in a coherent way, to present immigration authorities with a science base decisional method.

The immigration rules applying to an arriving passenger should of course reflect the ones of the highest RG country visited during the last few days, with the exception of transfer airports, which are considered safe.

## 7. Conclusions

In a pandemic such as Covid-19, opening the borders to tourism implies a certain amount of risk of importing the virus. Even vaccinations cannot guarantee total safety from virus spread, particularly since new variants show significant resistance. By using a selective entry policy based on a dynamic risk assessment (SEDRA) probability calculation algorithm such as the one proposed in this paper, we can minimize the number of imported cases while maximizing the income to the economy brought by visitors.

The SEDRA Project publishes bi-weekly up to date tables and charts on the following repository: https://sedraproject.github.io/index.html

## Data Availability

This study is based on the OWID Covid-19 public data published on: https://github.com/owid/covid-19-data/tree/master/public/data. The results of this study are updated every other week and made available on the SEDRA repository: https://sedraproject.github.io/index.html

## 9. Acknowledgements

The authors would like to thank Radhakhrishna Somanah for reviewing this work.

## 10. Funding

This project was carried out as a spontaneous volunteer initiative to help with the pandemic. No funding was requested or offered. Therefore there was no conflict of interest.

## APPENDIX A

Example of SEDRA table sorted by descending Safety Index

APPENDIX A : Example of SEDRA table sorted by descending Safety Index
| SEDRA report of 2022-06-30 |  |  |  |  |  |  |  |  |  | Sort: Descending Safety Index |  |
| --- | --- | --- | --- | --- | --- | --- | --- | --- | --- | --- | --- |
| COUNTRY | Safety Index | Risk Group | Probability infected | 1 UI every | Daily Deaths /Mio | Daily Tests /Mio | Imm. Level | Sev. Level | Conf. Level | Tot Cases /last 14 days | Population |
| Guinea | 100 | A: VERY LOW | 0.00E+00 | 0 | 0.00 | 0 | 10.00 | 0.00 | 0.01 | 0 | 13,497,237 |
| Ukraine | 100 | A: VERY LOW | 0.00E+00 | 0 | 0.00 | 0 | 10.00 | 0.00 | 0.00 | 0 | 43,466,822 |
| Tajikistan | 100 | A: VERY LOW | 0.00E+00 | 0 | 0.00 | 0 | 10.00 | 0.00 | 0.00 | 0 | 9,749,625 |
| Niger | 100 | A: VERY LOW | 0.00E+00 | 0 | 0.00 | 0 | 10.00 | 0.00 | 0.00 | 0 | 25,130,810 |
| Micronesia (country) | 100 | A: VERY LOW | 0.00E+00 | 0 | 0.00 | 0 | 10.00 | 0.00 | 0.00 | 0 | 116,255 |
| Gambia | 100 | A: VERY LOW | 0.00E+00 | 0 | 0.00 | 0 | 10.00 | 0.00 | 0.00 | 0 | 2,486,937 |
| Djibouti | 100 | A: VERY LOW | 0.00E+00 | 0 | 0.00 | 0 | 10.00 | 0.00 | 0.00 | 0 | 1,002,197 |
| Costa Rica | 100 | A: VERY LOW | 0.00E+00 | 0 | 0.00 | 0 | 10.00 | 0.00 | 0.00 | 0 | 5,139,053 |
| Congo | 100 | A: VERY LOW | 0.00E+00 | 0 | 0.00 | 0 | 10.00 | 0.00 | 0.00 | 0 | 5,657,017 |
| Central African Republic | 100 | A: VERY LOW | 0.00E+00 | 0 | 0.00 | 0 | 10.00 | 0.00 | 0.00 | 0 | 4,919,987 |
| Burkina Faso | 100 | A: VERY LOW | 0.00E+00 | 0 | 0.00 | 0 | 10.00 | 0.00 | 0.00 | 0 | 21,497,097 |
| Belarus | 100 | A: VERY LOW | 0.00E+00 | 0 | 0.00 | 0 | 10.00 | 0.00 | 0.00 | 0 | 9,442,867 |
| Egypt | 100 | A: VERY LOW | 0.00E+00 | 0 | 0.00 | 0 | 10.00 | 0.00 | 0.00 | 0 | 104,258,327 |
| Yemen | 90 | A: VERY LOW | 0.00E+00 | 0 | 0.00 | 0 | 9.03 | 0.00 | 0.00 | 0 | 30,490,639 |
| Chad | 82 | A: VERY LOW | 5.91E-08 | 16,914,985 | 0.00 | 0 | 8.17 | 0.00 | 0.00 | 1 | 16,914,985 |
| Liberia | 77 | A: VERY LOW | 7.72E-07 | 1,295,052 | 0.00 | 0 | 7.66 | 0.00 | 0.00 | 4 | 5,180,208 |
| Cambodia | 76 | A: VERY LOW | 1.00E-06 | 996,850 | 0.00 | 0 | 7.64 | 0.00 | 0.00 | 17 | 16,946,446 |
| Syria | 75 | A: VERY LOW | 3.83E-07 | 2,610,815 | 0.00 | 0 | 7.53 | 0.00 | 0.00 | 7 | 18,275,704 |
| Sierra Leone | 74 | A: VERY LOW | 2.46E-07 | 4,070,672 | 0.00 | 0 | 7.39 | 0.00 | 0.00 | 2 | 8,141,343 |
| China | 74 | A: VERY LOW | 3.72E-07 | 2,689,415 | 0.00 | 0 | 7.38 | 0.00 | 0.00 | 537 | 1,444,216,102 |
| Mali | 72 | A: VERY LOW | 5.75E-07 | 1,737,977 | 0.01 | 0 | 7.22 | 0.00 | 0.00 | 12 | 20,855,724 |
| Sudan | 71 | A: VERY LOW | 1.63E-06 | 615,197 | 0.00 | 0 | 7.07 | 0.00 | 0.00 | 73 | 44,909,351 |
| Myanmar | 71 | A: VERY LOW | 7.85E-07 | 1,274,558 | 0.00 | 0 | 7.06 | 0.00 | 0.01 | 43 | 54,806,014 |
| South Sudan | 70 | A: VERY LOW | 2.20E-06 | 455,255 | 0.00 | 0 | 7.03 | 0.00 | 0.00 | 25 | 11,381,377 |
| Tanzania | 70 | A: VERY LOW | 6.54E-06 | 152,981 | 0.00 | 0 | 6.99 | 0.00 | 0.00 | 402 | 61,498,438 |
| Algeria | 70 | A: VERY LOW | 1.28E-06 | 782,748 | 0.00 | 0 | 6.97 | 0.00 | 0.00 | 57 | 44,616,626 |
| Benin | 70 | A: VERY LOW | 7.55E-06 | 132,458 | 0.00 | 0 | 6.97 | 0.00 | 0.00 | 94 | 12,451,031 |
| Nigeria | 69 | A: VERY LOW | 1.57E-06 | 636,749 | 0.00 | 0 | 6.90 | 0.00 | 0.00 | 332 | 211,400,704 |
| Papua New Guinea | 68 | A: VERY LOW | 1.64E-06 | 607,934 | 0.00 | 0 | 6.76 | 0.00 | 0.00 | 15 | 9,119,005 |
| Cameroon | 67 | A: VERY LOW | 2.42E-06 | 412,489 | 0.01 | 0 | 6.66 | 0.00 | 0.00 | 66 | 27,224,262 |
| Kyrgyzstan | 66 | A: VERY LOW | 4.38E-06 | 228,564 | 0.00 | 0 | 6.62 | 0.00 | 0.00 | 29 | 6,628,347 |
| Somalia | 66 | A: VERY LOW | 3.36E-06 | 297,445 | 0.00 | 0 | 6.59 | 0.00 | 0.00 | 55 | 16,359,500 |
| Eritrea | 66 | A: VERY LOW | 3.05E-06 | 327,406 | 0.00 | 0 | 6.58 | 0.00 | 0.00 | 11 | 3,601,462 |
| Sri Lanka | 66 | A: VERY LOW | 2.70E-06 | 370,643 | 0.01 | 0 | 6.57 | 0.00 | 0.00 | 58 | 21,497,306 |
| Senegal | 66 | A: VERY LOW | 2.91E-06 | 343,926 | 0.00 | 0 | 6.57 | 0.00 | 0.00 | 50 | 17,196,308 |
| Libya | 65 | A: VERY LOW | 4.02E-06 | 248,519 | 0.00 | 0 | 6.52 | 0.00 | 0.00 | 28 | 6,958,538 |
| Nicaragua | 65 | A: VERY LOW | 1.06E-05 | 94,400 | 0.02 | 0 | 6.52 | 0.00 | 0.00 | 71 | 6,702,379 |
| Democratic Republic of Congo | 64 | A: VERY LOW | 0.00E+00 | 0 | 0.00 | 0 | 6.38 | 0.00 | 0.00 | 0 | 92,377,986 |
| Comoros | 63 | A: VERY LOW | 3.38E-06 | 296,152 | 0.00 | 0 | 6.33 | 0.00 | 0.00 | 3 | 888,456 |
| Timor | 63 | A: VERY LOW | 5.21E-06 | 191,982 | 0.00 | 0 | 6.33 | 0.00 | 0.00 | 7 | 1,343,875 |
| Nepal | 63 | A: VERY LOW | 6.13E-06 | 163,049 | 0.00 | 0 | 6.30 | 0.00 | 0.01 | 182 | 29,674,920 |
| Laos | 63 | A: VERY LOW | 7.3E-06 | 136,655 | 0 | 0 | 6.275 | 0 | 0 | 54 | 7379358 |
| Burundi | 63 | A: VERY LOW | 9.47E-06 | 105,650 | 0.00 | 0 | 6.26 | 0.00 | 0.00 | 116 | 12,255,429 |
| Guinea-Bissau | 62 | A: VERY LOW | 0.00E+00 | 0 | 0.00 | 0 | 6.22 | 0.00 | 0.01 | 0 | 2,015,490 |
| Malawi | 62 | A: VERY LOW | 8.04E-06 | 124,352 | 0.00 | 0 | 6.18 | 0.00 | 0.00 | 158 | 19,647,681 |
| Uganda | 61 | A: VERY LOW | 7.47E-06 | 133,874 | 0.01 | 0 | 6.10 | 0.00 | 0.00 | 352 | 47,123,533 |
| Togo | 61 | A: VERY LOW | 8.85E-06 | 113,043 | 0.02 | 0 | 6.07 | 0.00 | 0.00 | 75 | 8,478,242 |
| Pakistan | 61 | A: VERY LOW | 1.33E-05 | 75,142 | 0.01 | 0 | 6.06 | 0.00 | 0.00 | 2,997 | 225,199,929 |
| Afghanistan | 60 | A: VERY LOW | 1.14E-05 | 87,358 | 0.03 | 0 | 6.04 | 0.00 | 0.00 | 456 | 39,835,428 |
| Azerbaijan | 60 | A: VERY LOW | 3.52E-06 | 283,982 | 0.01 | 0 | 6.04 | 0.00 | 0.02 | 36 | 10,223,344 |
| Madagascar | 60 | A: VERY LOW | 1.31E-05 | 76,418 | 0.00 | 0 | 6.03 | 0.00 | 0.00 | 372 | 28,427,333 |

Abbreviation in this table: **UC**: Undetected Carrier, **Imm.**: Immunity, **Sev**: Severity, **Conf**: Confidence First page only inserted here. Full weekly reports on https://sedraproject.github.io/index.html

## APPENDIX B

Examples of Safety Index charts.

Note: The crammed look of country names is due to fitting the chart in a paper format. The online version is more readable as it allows you to isolate countries or view full screen. Full World charts on https://sedraproject.github.io/index.html

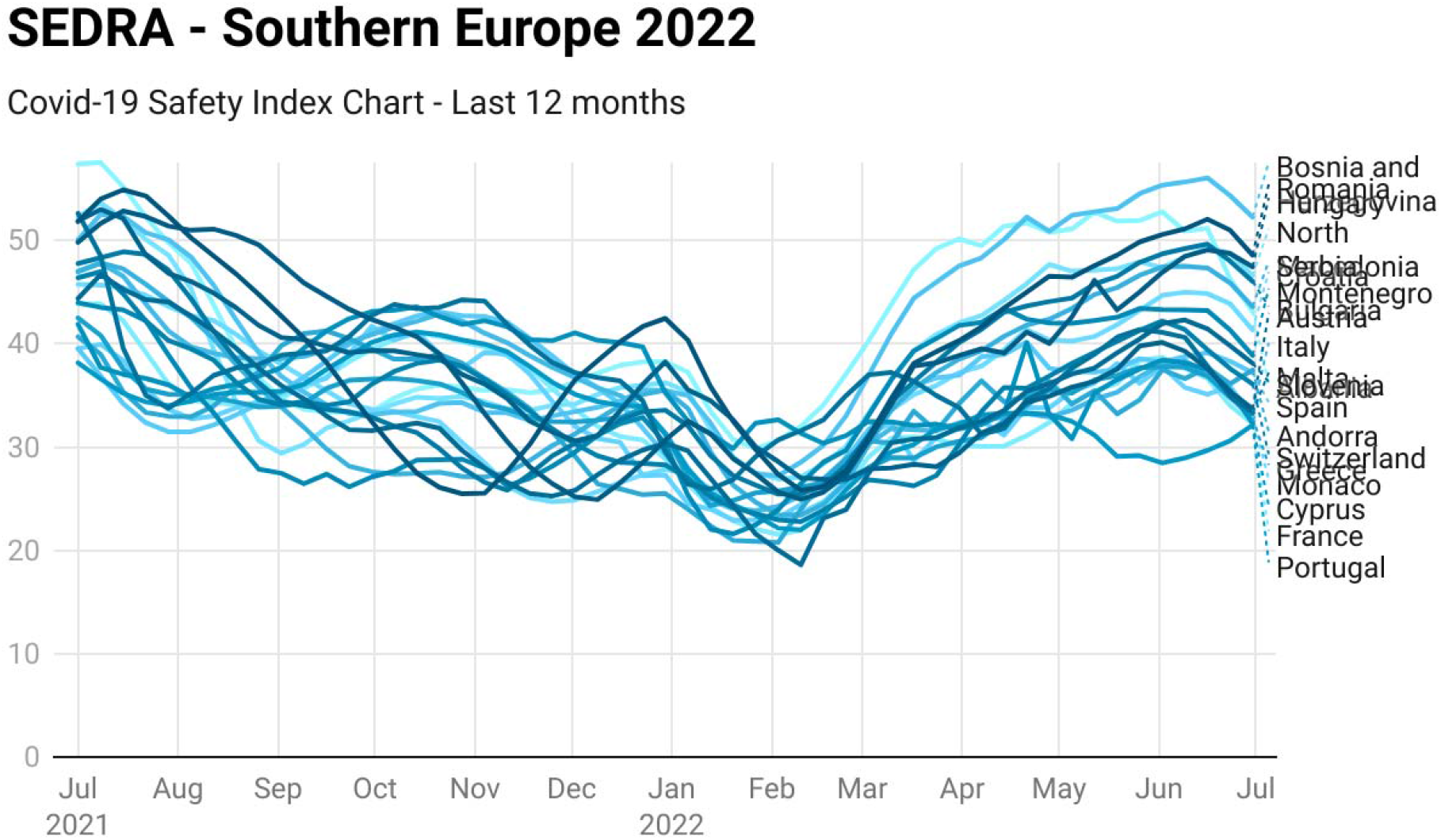

*The Safety Index reprints the probability that a passenger from a given country is Covid-free*

Chart: Silvano de Gennaro · Source: SEDRA Project · Created with Datawrapper

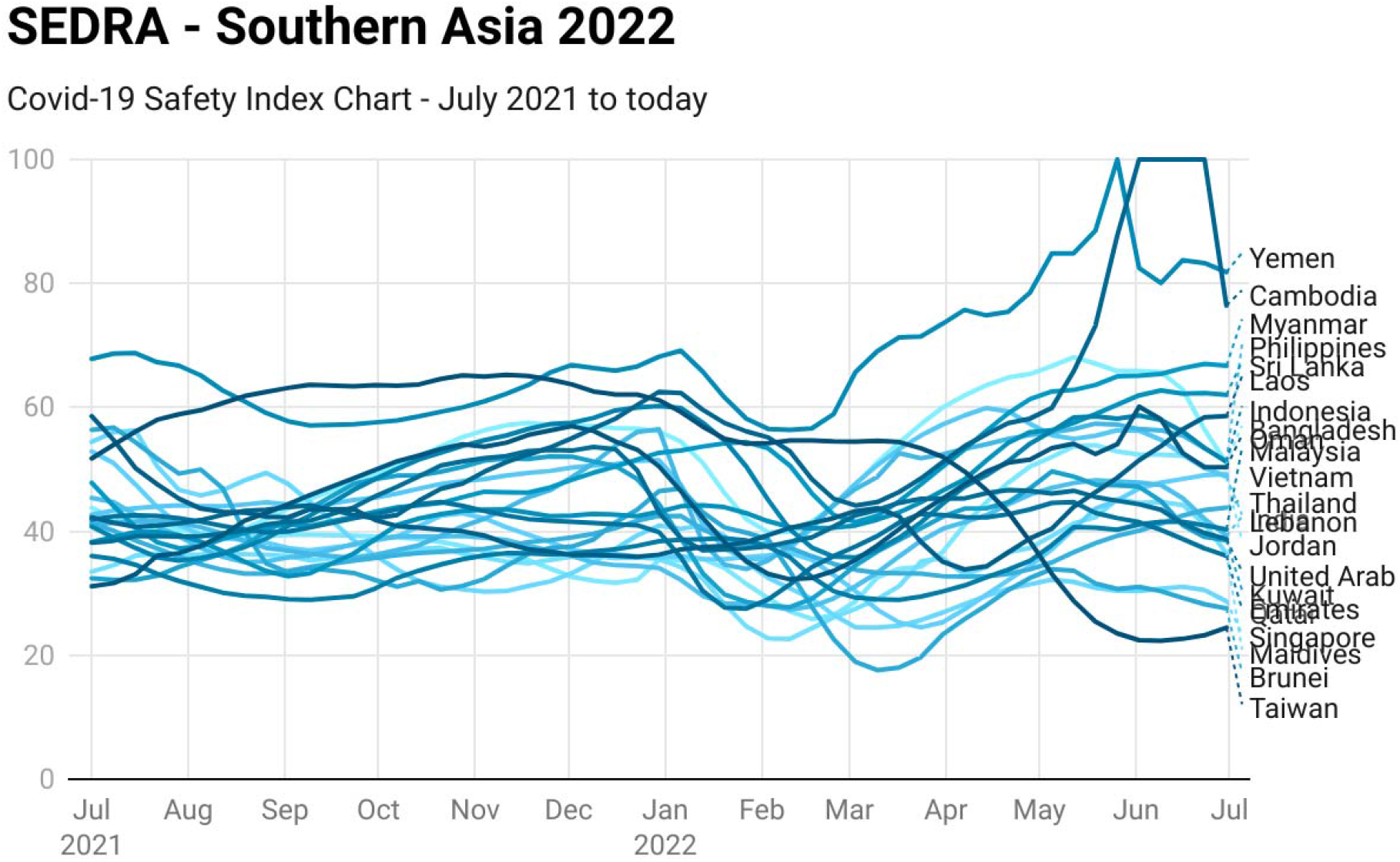

Chart: Silvano de Gennaro · Source: SEDRA Project · Created with Datawrapper

## Footnotes

1 Polymerase Chain Reaction

## Notes

### Competing Interest Statement

The authors have declared no competing interest.

### Funding Statement

This study did not receive any funding

### Summary of Updates

New Covid-19 variant Omicron (BA.1 - BA.5) involve different characteristics, in particular a shorter incubation time. The Safety-Index calculation formulas have been updated to reflect a variable incubation time, as opposed to a fixed 14 days associated to previous variants.

